# Building Prediction Models for 30-Day Readmissions Among ICU Patients Using Both Structured and Unstructured Data in Electronic Health Records

**DOI:** 10.1101/2021.08.10.21261858

**Authors:** Alex Moerschbacher, Zhe He

## Abstract

ICU readmissions are associated with poor outcomes for patients and poor performance of hospitals. Patients who are readmitted have an increased risk of in-hospital deaths; hospitals with a higher readmission rate have a reduced profitability, due to an increase in cost and reduced payments from Medicare and Medicaid programs. Predicting a patient’s likelihood of being readmitted to the ICU can help reduce early discharges, the risk of in-hospital deaths, and help increase profitability. In this study, we built and evaluated multiple machine learning models to predict 30-day readmission rates of ICU patients in the MIMIC-III database. We used both the structured data including demographics, laboratory tests, comorbidities, and unstructured discharge summaries as the predictors and evaluated different combinations of features. The best performing model in this study Logistic Regression achieved an AUROC of 75.7%. This study shows the potential of leveraging machine learning and deep learning for predicting ICU readmissions.

## 1 Introduction

Hospital readmission is defined as an episode when a patient who had been discharged from a hospital is admitted again within a specific time interval (“Hospital Readmission,” 2020). High readmission rates of patients are a signal of issues with healthcare quality [2], as high readmissions are indicators of persisting issues in a patient’s health (Ponzoni et al., 2017). The United State Centers for Medicare and Medicaid Services (CMS) includes hospital readmission rate as an important performance metric in their reimbursement decisions (“Hospital Readmission,” 2020). Yet one fifth of patients receiving Medicare benefits are readmitted within 30 days and 67% are readmitted within 90 days (Jencks et al., 2009). This outcome has both financial and medical consequences; the cost to Medicare of readmissions in 2004 was over $17 billion of avoidable costs. The high cost of avoidable readmission resulted in legislation which withholds a percentage of Medicare payments to hospitals with a high readmission rate (McIlvennan Colleen K. et al., 2015). During the first year of the Hospital Readmissions Reduction Program, 30% of hospitals received no penalty, 60% received a penalty less than 1% and 10% received the maximum penalty under the legislation of a withholding of 3% of total Medicare payments (McIlvennan Colleen K. et al., 2015). This resulted in total of $280 million in penalties (McIlvennan Colleen K. et al., 2015). Among patients who were readmitted, anywhere from 12% to 75% of these readmissions could have been avoided (Benbassat & Taragin, 2000). This high level of avoidable readmission indicates hospitals are not effectively utilizing preventative measures such as patient education, discharge assessments, and in-home aftercare options. This also presents great need of identifying patients who are more likely to be readmitted within 30 days so that hospitals can intervene as early as possible to avoid readmissions.

The wide adoption of Electronic Health Records (EHRs) presented an unprecedented opportunity for building machine learning and deep learning models to predict patient outcomes. EHRs contain fine-grained information about patient care including demographics, laboratory test results, medications, procedures, etc. The potential of using EHR for predicting 30 days readmission to ICU has been shown previously (Ben-Assuli & Padman, 2017; Futoma et al., 2015; Lin et al., 2019). These studies used demographic information, lab results, and chart events as predictors for machine learning models to predict if an individual will be readmitted to the ICU within 30-days. However, these studies have two major limitations: 1) They only used the structured data in EHRs (Futoma et al., 2015); and 2) they did not use deep learning for this problem, which may improve the prediction accuracy.

Most current studies only used demographic information, results from lab tests, chart events such as heart rate measurements and diagnoses on a patient’s chart. They did not consider how discharge notes, which contain rich information about patients and their care, could correlate to a patient’s likelihood of being readmitted to the ICU (Ben-Assuli & Padman, 2017; Futoma et al., 2015). For example, both Ben-Assuli and Padman (Ben-Assuli & Padman, 2017) and Futoma et al. (Futoma et al., 2015) limited the scope of their dataset to only the structured EHR. State-of-the-art language models that are pretrained with enormous amount of text corpora, such as the BERT model (Devlin et al., 2019), provide an opportunity to encode the semantic information from discharge notes for the machine learning models, while principal component analysis can reduce the dimensionality allowing for an even split between structured and unstructured data in the dataset.

Due to the huge number of parameters in deep learning models, it is time consuming to fine-tune the model to achieve optimal performance for a given dataset. Some prior studies, even though built deep learning models, did not seem to spend enough time for tuning the hyperparameters. For example, Lin et al. (Lin et al., 2019) did not mention hyperparameter tuning in their study while Futoma et al. (Futoma et al., 2015) performed minimal hyperparameter tuning in their study. Lin et al. (Lin et al., 2019) used a long short term memory to predict unplanned ICU readmission while Futoma et all (Futoma et al., 2015) gauged the ability of multiple deep neural networks to predict early hospital readmissions.

The aim of this study is to evaluate different machine learning and deep learning models using both the structured and unstructured data from the MIMIC-III dataset to predict if an individual will be readmitted to the ICU within 30 days of discharge. Structured data in this study include demographic, admission lab test data, and comorbidities (ICD-9-CM codes) for each patient. The unstructured data are the discharge notes for each patient and the respective embeddings generated from each discharge note. The contributions of this study are two-folds: 1) we evaluated the usefulness of unstructured data in EHRs for predicting ICU readmission rates; 2) we evaluated the effectiveness of hyperparameter tuning of the prediction models.

## 2 Methods

Figure 1 shows the overall workflow of the study. The structured data from the MIMIC-III database included the lab results, demographic information for each patient, and the admission data. ICD-9 codes were used to identify high level classification of comorbidities. Each patient’s recorded ICD9-CM codes in the MIMIC-III database was processed using the existing code (*GitHub - Jackwasey/Icd*, n.d.) to determine if the patient had any of the following conditions: myocardial infarction, congestive heart failure, peripheral vascular disease, cerebrovascular disease, dementia, chronic pulmonary disease, connective tissue disease-rheumatic disease, peptic ulcer disease, mild liver disease, diabetes without complications, diabetes with complications, paraplegia and hemiplegia, renal disease, cancer, moderate or severe liver disease, metastatic carcinoma, and HIV. This was then combined with the other structured data for each patient, nearest neighbor imputation was also performed for any missing values found in the lab test data for each patient.

**Figure 1:**
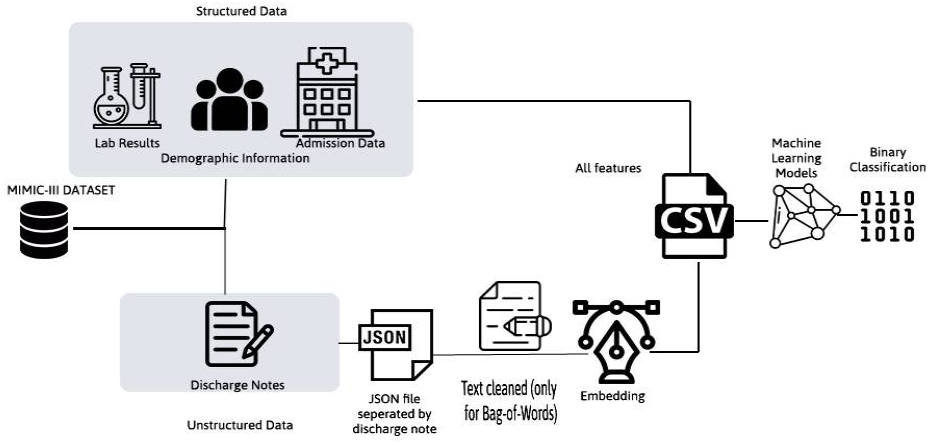
Overall Workflow of the Study

The unstructured data are the discharge notes for each patient. The unstructured data of discharge notes were in JSON files. Each discharge note was transformed into sentence embeddings using a transformer-based embedding method. In total six different embeddings were generated for each admission, this resulted in six different sentence embeddings for a single discharge note. The readmission notes were also cleaned to allow for a Bag-Of-Words approach to be used to represent the readmission notes. The Bag-Of-Words approach used the scikit package’s Count Vectorizer max feature parameter to limit the notes to the 3000 most frequently occurring words in the dataset. From there a matrix of token counts was created for these 3000 words. Principal component analysis was used to reduce the dimensionality of the unstructured data, so the unstructured data does not outnumber the structured data when combing the dataset. We created different feature sets using combinations of structured and unstructured data. Then we trained and evaluated prediction models to predict readmission for each admission.

### 2.1 Dataset

The Medical Information Mart for Intensive Care III (MIMIC-III) database was used in this study. MIMIC-III is a dataset of 40,000 patients who stayed in critical care units of Beth Israel Deaconess Medical Center between 2001 and 2012. The dataset contains demographics, vital sign measures taken on an hourly basis, laboratory test results, procedures, medications, free-text notes about the patients stay, and mortality reports.

Data selection was based on features found in similar studies that predict ICU readmission and mortality in the ICU (Ben-Assuli & Padman, 2017; Futoma et al., 2015). Patient dis-charge notes were also included in this study. Minimum, maximum, and average of lab features were included in this study to give a better view of the patient’s health throughout the course of their stay in the ICU. 47,388 admissions pertaining to 40,104 subjects from the database met the requirements of this study however only 4,522 were included. The limit was put in place to provide an equal split between individuals readmitted to the ICU and those who were not readmitted. The limiting factor was the number of individuals, 2,261, who were readmitted to the ICU. Table 1 provides the basic characteristics of the patient cohort. Table 2 shows the included features.

**Table 1:**
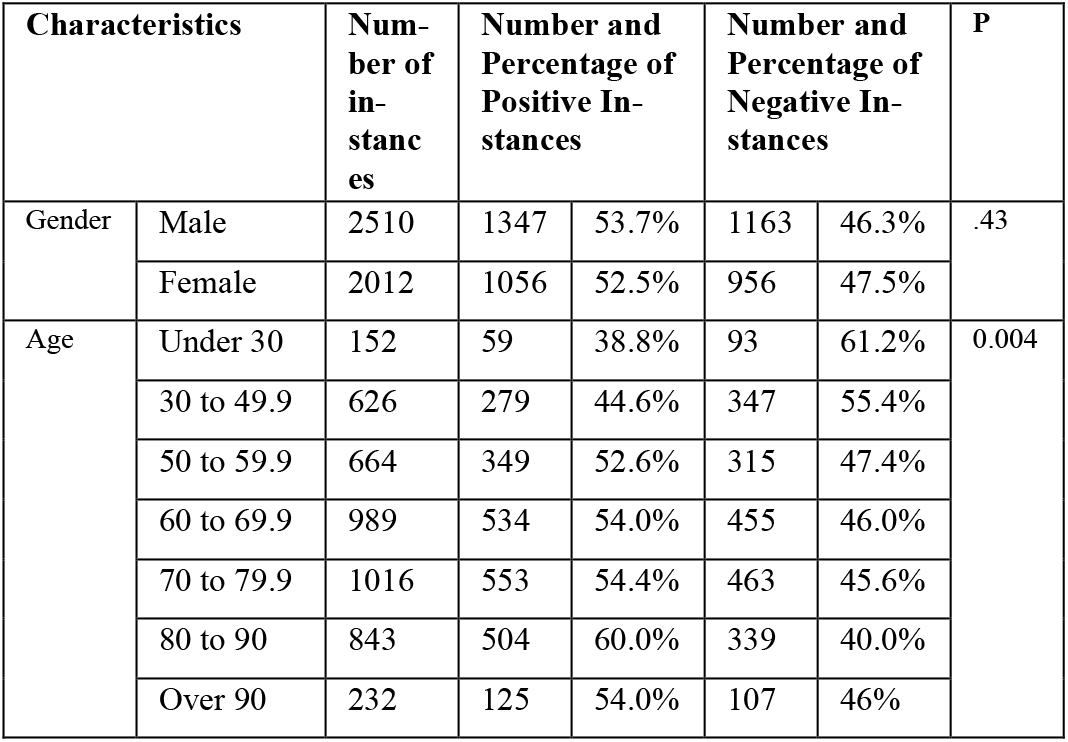
Basic characteristics of the patient cohort

| Characteristics |  | Number of instances | Number and Percentage of Positive Instances |  | Number and Percentage of Negative Instances |  | P |
| --- | --- | --- | --- | --- | --- | --- | --- |
| Gender | Male | 2510 | 1347 | 53.7% | 1163 | 46.3% | .43 |
|  | Female | 2012 | 1056 | 52.5% | 956 | 47.5% |  |
| Age | Under 30 | 152 | 59 | 38.8% | 93 | 61.2% | 0.004 |
|  | 30 to 49.9 | 626 | 279 | 44.6% | 347 | 55.4% |  |
|  | 50 to 59.9 | 664 | 349 | 52.6% | 315 | 47.4% |  |
|  | 60 to 69.9 | 989 | 534 | 54.0% | 455 | 46.0% |  |
|  | 70 to 79.9 | 1016 | 553 | 54.4% | 463 | 45.6% |  |
|  | 80 to 90 | 843 | 504 | 60.0% | 339 | 40.0% |  |
|  | Over 90 | 232 | 125 | 54.0% | 107 | 46% |  |

**Table 2:**
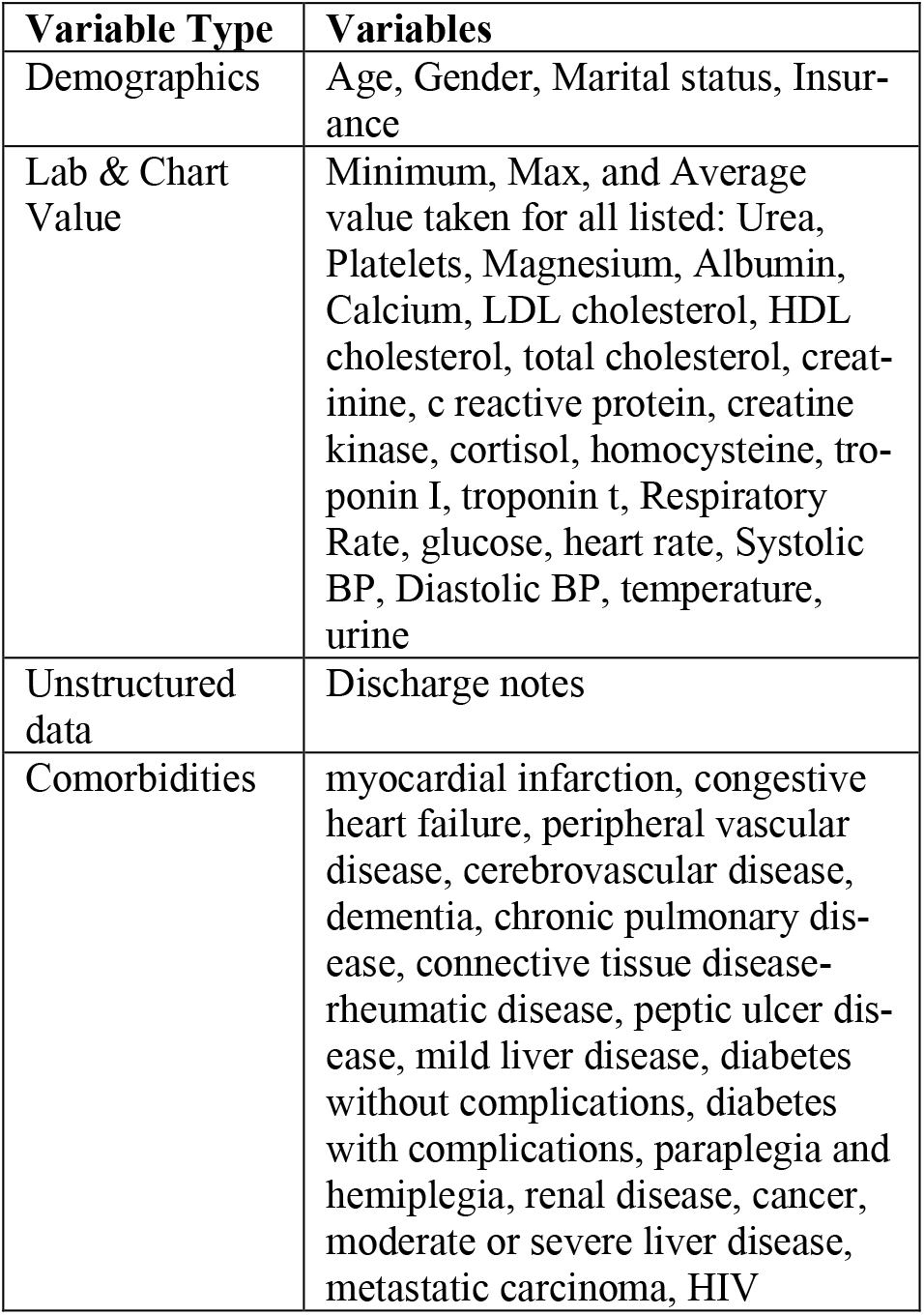
Structured and unstructured data from MIMIC-III

### 2.2 Data Preprocessing

Nearest neighbor imputation was employed to impute missing lab results. These missing results may be because some patients did not have those lab tests or these values were not accurately reported. Categorical variables with a high degree of cardinality, anything greater than 10, were dropped from the dataset.

The Sentence-BERT base model was used to generate sentence tokens from the discharge notes (Reimers & Gurevych, 2019). Sentence-BERT base model is a transformer model that uses Siamese and triplet network structures to generate sentence embeddings (Reimers & Gurevych, 2019). This model is an improvement of the BERT base model as it maintains the same level of accuracy while reducing the computation time dramatically. Six additional BERT-based models were also used to generate sentence transformers stsb-distilbert-base, bert-base-nli-max-tokens, bert-base-nli-mean-tokens, distilbert-base-nli-stsb-mean-tokens, stsb-roberta-large, and roberta-base-nli-stsb-mean-tokens (Wolf et al., 2020). After generating the sentence embeddings and transformers, principal component analysis was used on the sentence embeddings, to reduce the dimensionality to match the number of features in the structured dataset. As such, the combined structured data and unstructured data will be transformed to a vector that includes the same amount of features from the structured data and the unstructured data. Before performing PCA, the embeddings produced 767 features for each record. PCA was then used to reduce the number of features to 50 to create a balance between the representation of structured and unstructured data in the dataset. When the comparison of only unstructured data was being performed, PCA was not used.

To generate the binary 30-day readmission outcome variable, we compared the ICU discharge date with their next admission date to determine if readmission occurred within 30 days. If the difference between the two dates was within 30 days, the readmission variable was set to true for that admission.

### 2.3 Modeling and Evaluation

The outcome of the prediction model is to predict if an admission will be followed by a readmission in 30 days. Three datasets were created: (1) one containing only structured data, (2) one containing unstructured data and its respective embeddings, and (3) a combined dataset of structured and unstructured data. The following popular classification algorithms were used: Logistic Regression, XGBOOST, Random Forest, Feed Forward Neural Network, and Support Vector Classification. We used standard metrics including accuracy, precision, recall, and area under the curve to evaluate their performance. Scikit-learn was used to implement each of the classification algorithms (Pedregosa et al., n.d.). We randomly split the instances into 80% for training and 20% for testing.

A Random Forest classifier is made up of a group of decision trees with each tree being as unrelated to the other trees as possible. This is accomplished by using bagging and feature randomness when creating each tree to minimize the correlation chance. Once the trees are created each individual tree in the set returns a class prediction, the class with the most “votes” by the trees is the model’s prediction. The underlying principle guiding this algorithm is that a group of highly unrelated models will have a better outcome than any single model. We finetuned the parameters of this model by first starting out with the default hyperparameters. From there the criterion was changed between gini and entropy and the number of trees were increased by an interval of 500 in the model until the performance of the model decreased.

Rather than training the models in isolation from one another, boosting trains each model in sequential order with the next model fixing the errors of the former. This process continues until no further improvements can be made in the model. This prevents the issue of Random Forest where uncorrelated models make the same mistakes. XGBOOST is a model which is designed to support the idea of additive tree model which optimizing the process to be time effective (Chen & Guestrin, 2016). We finetuned the parameters of this model with all datasets having using hinge loss for binary classification and increasing the number of tress in the model until performance of the model decreased.

Feedforward neural networks are deep learning model used when the data is not sequential or time dependent. As the flow of data in the model only moves towards the output in comparison with recurrent neural networks where the data is cyclical. This deep learning model works for our scenario as each admission is separate from other admissions in the dataset, and the data in the dataset is not time series data. We finetuned the hyperparameters for each dataset. The combined dataset used a model with 100 epochs and 3 hidden layers with 150, 100, 50 neurons respectively, an activation function of relu, and a solver of adam. The structured dataset used a model with 300 iterations. The unstructured dataset used a max iteration of 10000, 8 hidden layers, with 100 neurons in each layer, and an alpha of 0.003 and early stopping enabled. The increase in iterations, hidden layers in the unstructured dataset can be explained by the difference in features. In the unstructured dataset there were 767 features while in the combined dataset there were 100 features and the structured dataset there were 50 features. For any parameters not specified here, we used scikit’s default parameters for the models.

A Support Vector Classification Algorithm attempts to find a hyperplane in an N-dimensional space (N – number of features) that can classify the data points. Overall objective is to find a hyperplane that results in the greatest distance between data points in both classes. The easiest way to visualize this is the hyperplane is a dividing line between two classification groups with the line giving each group the greatest distance from another. As the number of features in a dataset increases so does the dimensionality of the hyperplane, making visualization difficult as the feature set increases. The position of new data points in relation to line results in them data’s classification by the model. For the SVC model we used the default parameters for the structured dataset and increased the number of iterations to 1050 for the combined and unstructured dataset.

## 3 Results

Table 3 shows the performance of the models using the structured data only. The highest scoring model in the structured dataset was the Random Forest model with an accuracy of 61.1%, a recall of 81.3%, a precision score of 73.4%, an F1 score of 77.2%, and an AUROC of 73.9%. This model has the second highest AUROC score out of all three datasets.

**Table 3:**
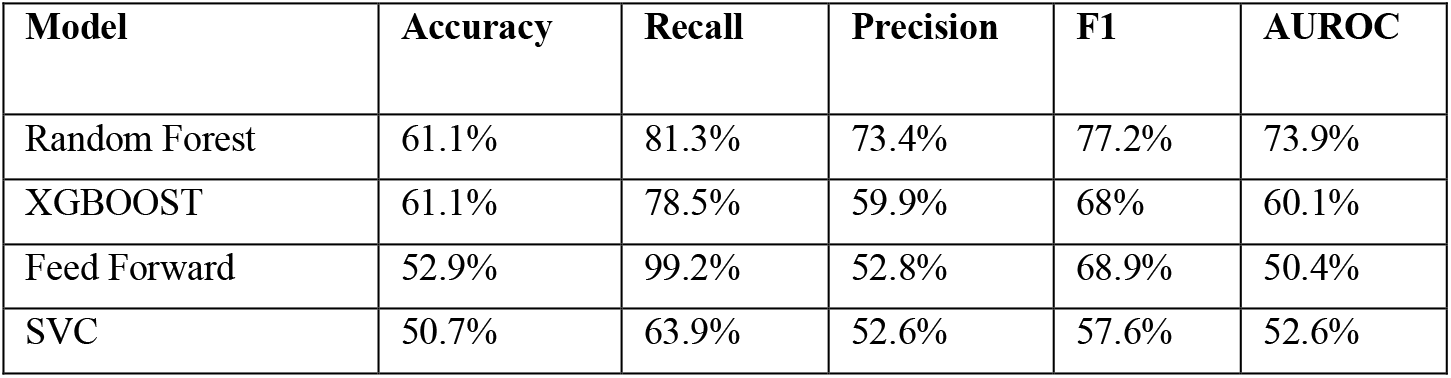
Performance of the models using structured data

| Model | Accuracy | Recall | Precision | F1 | AUROC |
| --- | --- | --- | --- | --- | --- |
| Random Forest | 61.1% | 81.3% | 73.4% | 77.2% | 73.9% |
| XGBOOST | 61.1% | 78.5% | 59.9% | 68% | 60.1% |
| Feed Forward | 52.9% | 99.2% | 52.8% | 68.9% | 50.4% |
| SVC | 50.7% | 63.9% | 52.6% | 57.6% | 52.6% |

Table 4 shows the performance of the models using the unstructured dataset. The Logistic Regression model was the best overall performing model. 68.5% of all predictions were correct, and the model achieved an AUROC score of 75.7%. The recall rate of 68.2% for this model indicates the Logistic Regression model was able to correctly identify 68.2% of all patients who would be readmitted to the ICU within 30 days. The model also achieved an F1 score of 69.7%.

**Table 4:** Performance of the models using unstructured data (Top 8 Models)

| Model | Embedding | Accuracy | Recall | Precision | F1 | AUROC |
| --- | --- | --- | --- | --- | --- | --- |
| Logistic Regression | Bag-Of-Words | 68.5% | 68.2% | 71.2% | 69.7% | 75.7% |
| Random Forest | Bag-Of-Words | 65.8% | 76.5% | 65.7% | 70.7% | 64.9% |
| SVC | Bag-Of-Words | 64.8% | 69.9% | 66.5% | 68.2% | 64.4% |
| Feed Forward | Bag-Of-Words | 63.5% | 68.6% | 65.4% | 66.9% | 63.0% |
| XGBOOST | Bag-Of-Words | 63.3% | 76.8% | 63.1% | 69.3% | 62.2% |
| Feed Forward | bert-base-nli-mean-tokens | 60.8% | 64.3% | 64.6% | 64.5% | 60.5% |
| Random Forest | bert-base-nli-mean-tokens | 60.1% | 69.9% | 62.3% | 66.2% | 59.6% |
| SVC | bert-base-nli-mean-tokens | 62.9% | 70.1% | 62.4% | 66.0% | 59.1% |

Table 5 shows the performance of the models using the combined dataset. The Random Forest model was the best overall performing model in the combined dataset. 70.8% of all predictions were correct, and the model achieved an AUROC score of 70.3%. The recall rate of 79.2% for this model indicates the Random Forest model was able correctly identify 79.2% of all patients who would be readmitted to the ICU within 30 days. The model also achieved an F1 score of 74.0%. The only models with a higher recall score in the combined dataset were SVC and XGBOOST models, however they are not displayed due to their low AUROC score. The discussion section will provide an explanation for why the higher AUROC score is occurring.

**Table 5:** Performance of the models using combined data (Top 8 Models)

| Model | Embedding | PCA | Accuracy | Recall | Precision | F1 | ROC |
| --- | --- | --- | --- | --- | --- | --- | --- |
| Random Forest | stsb-distilbert-base | 50 | 70.8% | 79.2% | 69.6% | 74.1% | 70.4% |
| Random Forest | Bag-Of-Words | None | 66.7% | 75.3% | 63.6% | 69.0% | 66.8% |
| SVC | Bag-Of-Words | None | 66.5% | 67.0% | 65.6% | 66.3% | 66.5% |
| XGBOOST | Bag-Of-Words | None | 64.2% | 70.6% | 61.9% | 66.0% | 64.3% |
| Feed Forward | Bag-Of-Words | None | 62.1% | 61.6% | 61.4% | 61.5% | 62.1% |
| Random Forest | roberta-base-nli-stsb-mean-tokens | 50 | 62.1% | 73.7% | 61.7% | 67.1% | 61.5% |
| Random Forest | stsb-roberta-large | 50 | 61.9% | 73.3% | 61.6% | 66.9% | 61.3% |
| Random Forest | bert-base-nli-max-tokens | 50 | 61.8% | 74.2% | 61.4% | 67.2% | 61.2% |

On average the model performance in the unstructured dataset was lower than both the combined and structured dataset. The next highest scoring machine learning model which did not use the bag-of-words approach for the unstructured data was a Feed Forward model which used bert-base-nli-mean-tokens for its embeddings. This model had an ROC score of 60.5%.

## 4 Discussion

### 4.1 Principal Results

This study compared multiple different machine learning models for predicting ICU readmission within 30 days of patients using the MIMIC-III dataset. In our dataset, the deep learning models consistently underperformed non-deep learning models. Overall, the Random Forest model consistently outperformed all other machine learning models with an AUROC of 73.9% in the structured dataset and an AU-ROC of 70.3% in the combined dataset. Logistic Regression was the only model able to outperform the random forest model with an AUROC score of 75.7% in the unstructured dataset. Embedding choice did not have a major impact on the AUROC score of a model. The average difference of each model between their highest and lowest scoring embedding was 4.15%. The feed forward neural network model has the highest difference with a 7% difference in AUROC score. While the Random Forest model had the lowest difference of 2.2% between the highest and lowest AUROC score for each embedding. Hyperparameter tuning did result in an overall improvement in deep learning models. While some models such as the Random Forest could have the same parameters applied across datasets with improved outcomes, both deep learning models required hyperparameter tuning specific to each dataset. This is especially apparent in the feed forward neural network model which required 10000 maximum iterations in the combined dataset but only 300 in the structured dataset. Due to the complexity of the deep learning models and the requirement to retrain the model after changing a single parameter a significant amount of time is required to tune parameters for deep learning models.

When comparing the results of this study with other studies the highest performing model was outperformed by Lin et al. (Lin et al., 2019) which had an AUROC of 79.1% while our model had an AUROC of 75.7%. The difference in results in the Lin et al. study can be explained by their use of time series data which provided the deep learning models in their study additional data. This study did not use time series data, which is challenging to construct and may suffer from the granularity issue for different variables (Cirillo et al., 2021).

In the combined dataset, the SVC and XGBOOST models were able to outperform the other models when comparing their recall scores. While the SVC and XGBOOST model performed substantially lower in comparison in other evaluation areas. By showing the prediction for each admission in the SVC and XGBOOST models, it is apparent why their recall scores are so high. Both models were more likely to predict that a patient would be readmitted to the hospital. Since nearly all predictions were positive the models had a high recall score but an overall low score in other metrics due to the high number of false positives in the balanced dataset.

### 4.2 Limitations and Opportunities

A few limitations should be noted. Imputation had to be used for some lab values because not all lab values are collected for all the patients. According to our recent paper (Payrovnaziri et al., 2020), imputation techniques may impact the stability of the prediction performance and the feature ranking results.

Similar with other machine learning projects, the process of manually tuning parameters was extremely time-consuming since the model would have to be retrained each time a pa-rameter was changed to gauge its performance. The complexity of the models resulted in a long training time. Future studies should look into automating the tuning process such as grid search. Other future direction is to apply post hoc interpretability enhancement method to explain how the models work and why a certain prediction is made.

## 5 Conclusion

In this study, we predicted unplanned ICU readmission using three different datasets, a structured dataset with chart events, lab results, and demographic information, an unstructured dataset with the sentence embeddings for each admission discharge notes using multiple encoders, and a combined dataset composed of both the structured and unstructured data. The results of this study showed that the Logistic Regression model using Bag-Of-Words embedding had an AUROC of 75.7% and recall of 68.2% using only the unstructured dataset.

## Data Availability

The MIMIC III data used in this study is publicly available.

https://mimic.mit.edu/

## Acknowledgments

This study was partially supported by the National Institute on Aging (NIA) of the National Institutes of Health (NIH) under Award Number R21AG061431; and in part by Florida State University-University of Florida Clinical and Translational Science Award funded by National Center for Advancing Translational Sciences under Award Number UL1TR001427.

